# The mechanical and perfusion basis of exercise limitation in apical hypertrophic cardiomyopathy

**DOI:** 10.1101/2023.07.14.23292698

**Authors:** Rebecca K. Hughes, James W. Malcolmson, Ricardo P. Monteiro, Camilla Torlasco, Shafik Khoury, Peter Kellman, Hui Xue, Rhodri Davies, Charlotte Manisty, Thomas A. Treibel, Luis R. Lopes, Saidi A. Mohiddin, Gabriella Captur, James C. Moon, Guy Lloyd

**Affiliations:** Institute of Cardiovascular Science, University College London, Gower Street, London, UK; Barts Heart Centre, Department of Cardiac Diagnostics and The Inherited Cardiovascular Diseases Unit, St Bartholomew’s Hospital, West Smithfield, London, UK; Centre for Advanced Imaging. William Harvey institute, Queen Mary University of London, UK; Faculty of Medicine and Biomedical Science, University of Algarve, Faro, Portugal; Department of Cardiology, IRCCS Istituto Auxologico Italiano, Milan, Italy; Cardiovascular Clinical and Academic Group, Molecular and Clinical Sciences Institute, St. George’s University of London, London, United Kingdom; National Heart, Lung, and Blood Institute, National Institutes of Health, DHHS, Bethesda, Maryland, USA; Inherited Heart Muscle Conditions Clinic, Gower Street, Department of Cardiology, Royal Free Hospital, NHS Trust, UK; University College London MRC Unit of Lifelong Health and Ageing, 1-19 Torrington Place, Fitzrovia, London, UK

**Keywords:** Apical hypertrophic cardiomyopathy, Cardiac magnetic resonance imaging, Transthoracic echocardiography, Exercise echocardiography, Cardiopulmonary exercise testing, Multimodality imaging

## Abstract

**Background:** Apical hypertrophic cardiomyopathy (ApHCM) patients can develop symptoms (chest pain, breathlessness), cardiac structural abnormalities (atrial dilatation, scar, apical aneurysm) and adverse outcomes despite preserved systolic function. Underlying mechanisms are poorly understood. We hypothesized that functional limitation in ApHCM may be associated with altered myocardial mechanics and myocardial perfusion.

**Methods:** We recruited 42 ApHCM patients and compared them with healthy controls (n=36). We assessed functional limitation (VO_2_ <80% predicted) using cardiopulmonary exercise testing, stress apical myocardial blood flow (MBF) and scar using cardiovascular magnetic resonance, and echocardiography global longitudinal strain (GLS) and twist at rest and during exercise.

**Results:** Functional limitation occurred in 35% vs 6% of controls (*P*<0.005) and was unrelated to wall thickness or ejection fraction. Myocardial mechanics were abnormal, with impaired GLS (−11.0% vs −18.3%, P<0.001), increased LV twist (22.6±9⸰ vs 16.6±4⸰, P<0.005) and delayed diastolic untwist (17.9% vs 9.2% of diastole, P<0.005). With exercise, GLS, twist and twist rate augmented but diastolic untwist delayed further. Stress apical MBF was reduced in all ApHCM patients and associated with mechanical abnormalities (GLS P<0.001, delayed diastolic untwist P=0.039). Percentage predicted peak VO_2_ was worse with lower apical blood flow (P<0.005) and reduced GLS (P=0.017), but the best predictor was prolonged diastolic untwist (β-0.828, P<0.05).

**Conclusion:** One third of ApHCM patients have functional limitation - best predicted by delayed diastolic untwist. GLS, twist mechanics and apical MBF were abnormal in all subjects highlighting mechanical and perfusion abnormalities as hallmarks of the disease, but identifying diastolic impairment as the mechanistic link.

**Delayed myocardial untwist predicts functional limitation and is linked to microvascular ischemia in Apical HCM:** 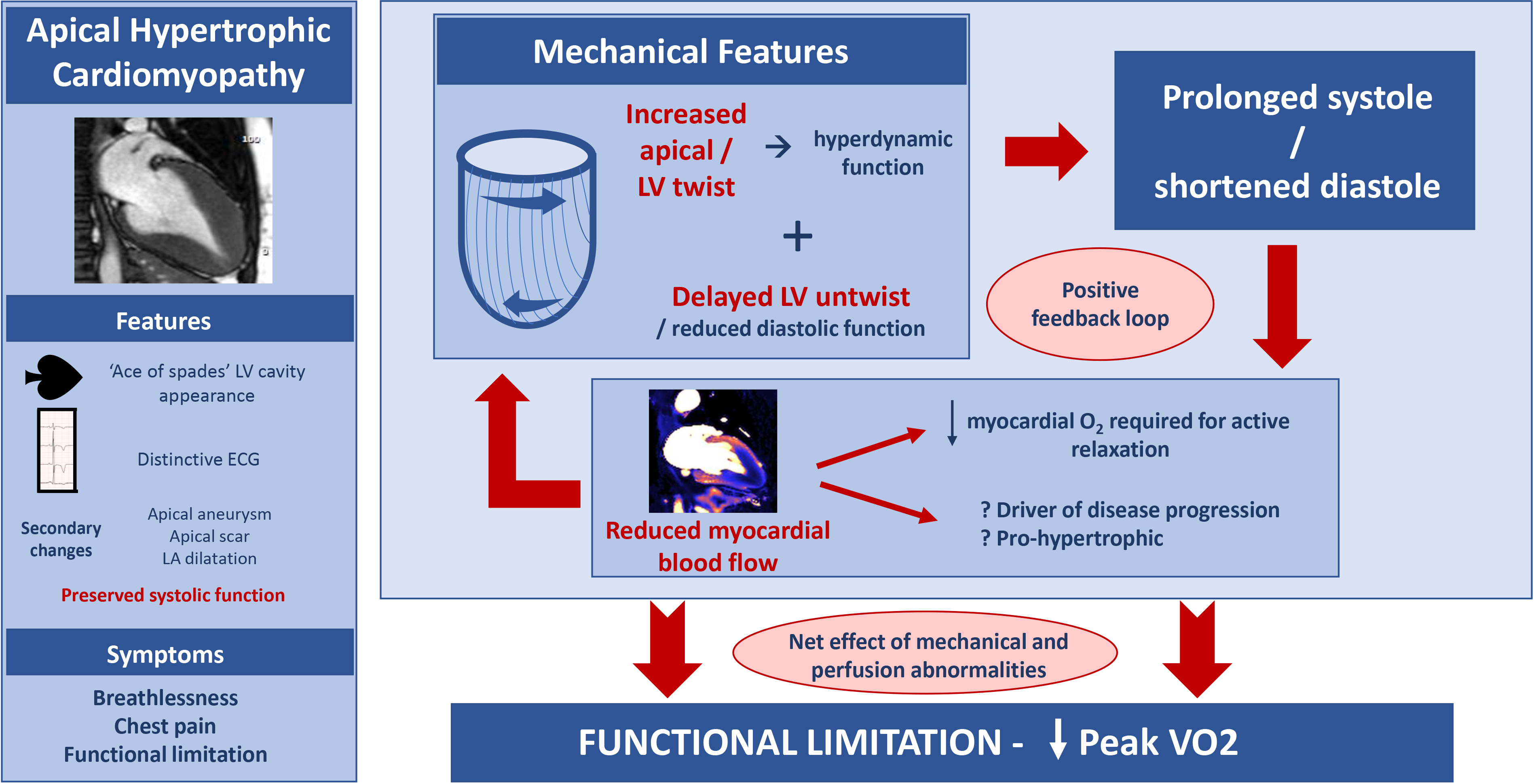

We postulate that increased left ventricular (LV) twist and delayed diastolic untwist results in prolonged systole and shortened diastole, which in turn reduces myocardial blood flow in a positive feedback loop. The net effect of mechanical and perfusion abnormalities is of functional limitation.

**CLINICAL PERSPECTIVES:** Apical hypertrophic cardiomyopathy (ApHCM) patients can develop symptoms, functional limitation, and adverse outcomes but the mechanisms underpinning this are unknown. Functional limitation is best assessed using cardiopulmonary exercise testing measuring peak VO_2_, whereby a value <80% predicted (based on age, sex and body size) is abnormal. Peak VO_2_ is a known prognostic measure in hypertrophic cardiomyopathy. This multi-modality imaging study aimed to explore whether functional limitation associated with abnormal myocardial mechanics and myocardial perfusion. We found that 35% of ApHCM patients had functional limitation (vs 6% healthy controls), which was independent of wall thickness and ejection fraction. Percentage predicted peak VO_2_ was worse with lower apical myocardial blood flow and reduced GLS, but was predicted by delayed diastolic untwist, implicating diastolic impairment as the mechanistic link. Understanding the abnormal mechano-structural and physiological features that contribute to, or predict functional limitation in apical hypertrophic cardiomyopathy strengthens our understanding of the disease and provides focus for future targeted research.

## INTRODUCTION

Apical hypertrophic cardiomyopathy (ApHCM) accounts for up to 25% of cases of hypertrophic cardiomyopathy (HCM) and is characterised by apical left ventricular (LV) hypertrophy (LVH), systolic obliteration of the apical LV cavity and anterior T-wave inversion. There are several key differences between ApHCM and other morphological subtypes; these include a lower prevalence of identifiable genetic mutations, distinctive and omnipresent electrocardiogram (ECG) features (tall R-waves and deep T-wave inversion) and a different risk profile (higher prevalence of atrial fibrillation (AF) and stroke; similar mortality rates)^1^. Left atrial (LA) dilatation is common. HCM is a disease marked by heterogeny; the causes and consequences of the fidelity between ApHCM’s associated features, and ApHCM’s distinctiveness from other forms of HCM are not well understood.

Conventionally, a wall thickness in the LV apex that exceeds 15mm has been required for diagnosis, but morphologically milder disease, termed ‘relative’ ApHCM, is increasingly recognised (typical ECG, apical systolic cavity obliteration, but with wall thickness <15mm^2^. In some cases, the disease may progress to include scarred, akinetic apical aneurysms, which have been associated with adverse prognosis^3, 4^. Symptoms are common, with chest pain and dyspnoea the most reported.

Cardiac imaging demonstrates multiple functional abnormalities in ApHCM. Apical hypertrophy causes the familiar “spade-like” cavity and results in apical cavity obliteration early in systole and persisting well after aortic valve closure, generating high pressures and creating basal to apical heterogeny in myocardial mechanics across the cardiac cycle, including regional differences in myocardial deformation and diastolic function^5, 6^. Conventional measures of systolic function, such as ejection fraction (EF) may be supranormal, but more advanced echocardiography derived parameters may be very abnormal either globally (global longitudinal strain(GLS)^7–9^, or apically (longitudinal, radial and circumferential strain, including twist)^9–11^. Furthermore, cardiovascular magnetic resonance (CMR) using fully quantitative perfusion mapping demonstrated that apical perfusion defects are a universal feature in ApHCM, occurring across the phenotypic spectrum, including those with relative ApHCM^12^. Notably, early data suggests that regional differences in myocardial mechanics and blood flow may be causally related^6^.

Reduced exercise capacity in HCM is widely reported and can be objectively measured with peak oxygen uptake (peak VO_2_) using cardiopulmonary exercise testing (CPEX), which has prognostic significance^13^. In non-apical HCM, GLS increases during exercise, but twist does not, suggesting a lack of exercise reserve^13^. The duration of time for the LV to untwist in diastole has also been shown to predict peak VO_2_ ^14^. In ApHCM, functional limitation has been little explored, and the underpinning roles for abnormal myocardial mechanics and perfusion abnormalities are unknown. We hypothesized that functional limitation in ApHCM is common and independent from conventional measures of phenotypic severity in HCM; instead, we sought to determine if abnormalities in global/regional myocardial mechanics (strain, twist) and in global/regional myocardial blood flow were associated with objective limitations in functional capacity.

## METHODS

A prospective study approved by the National Health Service Research Ethics Committee (NHS REC) and Health Research Authority (HRA) and conducted in accordance with the Declaration of Helsinki. All subjects provided written, informed consent (REC 18/LO/0188 and 15/LO/0086).

### Study Population

We prospectively recruited patients with ApHCM from tertiary referral cardiomyopathy clinics at St Bartholomew’s Hospital and St George’s Hospital, London, UK. They were subclassified based on current diagnostic criteria as ‘overt ApHCM’ (defined as maximum apical wall thickness ≥15mm in end-diastole) and relative ApHCM (defined previously^2^ as inappropriate apical hypertrophy compared to expected apical wall thickness (loss of apical tapering, apical thickness>basal thickness) but <15mm, and other characteristic features of the disease (distinctive ECG changes^15^, apical cavity obliteration^6^ or apical aneurysm)). Other inclusion criteria were 1) age ≥18, 2) no secondary causes of LVH, 3) no known coronary artery disease. Healthy volunteer (HV) controls underwent exercise echocardiography and CPEX and had no significant past medical history, including cardiovascular disease. Exclusion criteria were those unable to exercise, the presence of conventional contraindications to CMR and those with permanent pacemakers/internal cardiac defibrillators.

### Exercise echocardiography and cardiopulmonary exercise test acquisition

Transthoracic echocardiography (TTE - Vivid E95, GE Medical System, Horten, Norway) was performed using a standard 2D probe. The protocol consisted of apical 4-, 3- and 2-chamber views, focused LV Q-stress (colour tissue Doppler imaging) in the apical 4-chamber view, continue wave doppler (CWD) imaging through mitral valve, mid-LV cavity, left ventricular outflow tract (LVOT) and tricuspid valve. Speckle tracking echocardiography was performed with a frame rate of 50-80 frames/second and a probe frequency of 1.7-2.0 MHz. Images were optimised for sector width and image depth to maintain adequate frame rate without compromising image quality^16^. Focused apical 4-, 3- and 2-chamber views were obtained to derive longitudinal strain. Parasternal short axis views at the level of the mitral valve (defined as the point of the tips of the mitral valve leaflets) and apex (defined as the apical-most level visible). As cavity obliteration is a key feature of the disease, we accepted LV luminal obliteration at this level to represent the true apex. Images were acquired and stored over a single cycle and were acquired at baseline (rest) and during early exercise (intermediate intensity, defined as a heart rate rise of 40-80% increase in heart rate, ∼100bpm).

CPEX was simultaneously performed on a semi-recumbent bicycle (ERG 9/11 SL, Schiller, Switzerland) using exercise protocols determined on an individual basis by the operator based on functional status and reported exercise capacity of each individual subject (5, 10, 15 or 20W ramp protocol). Work rate increased linearly during the exercise phase of the test until voluntary exhaustion aiming for a test duration of 8-12 minutes. Continuous 4-lead ECG tracing was monitored throughout the test, noting any dynamic ST or T wave changes and arrhythmias (the latter being an indication to abort the test). CPEX parameters were collected using a CPEX mask attached to calibrated oxygen cylinder and a sampling line collecting and recording expiratory gases on a breath-by-breath basis and analysed using a respiratory cart (Schiller CS 200, Switzerland or Quark, Cosmed, Italy). Blood pressure measurements were recorded at rest and at regular intervals during the test. At the start of each test, a 2-minute rest period was used to collect respiratory gas exchange and haemodynamic measurements. During exercise, subjects were asked to cycle and maintain 60-70 rpm throughout the test, and to continue pedalling until they reached a cause for test cessation (maximum workload, achieved target heart rate, exercise limitation) or if any abnormal test criteria were met (intolerable symptoms, muscular exhaustion, symptomatic hypotension, arrhythmias, significant hypertension).

### CMR acquisition

CMR scans were performed as previously described^17^ on a 1.5 Tesla magnet (Aera, Siemens Healthcare, Erlangen, Germany) using a standard clinical protocol consisting of cine imaging, stress and rest perfusion mapping and late gadolinium enhancement (LGE) with automated motion correction and phase-sensitive inversion recovery. Vasodilator stress perfusion used adenosine (140 mcg/kg/min for 4 minutes, increased to 175 microgram/kg/min for a further 2 minutes if less than 10 bpm heart rate increase or no symptoms). A gadolinium-based contrast agent (gadoterate meglumine, Dotarem, Guerbet, Paris, France) was injected into a peripheral vein during peak vasodilator stress at 0.05mmol/kg. 60 images were typically acquired for basal, mid and apical left ventricular (LV) short-axis slices. Rest perfusion images were subsequently acquired after 6-10 minutes. Perfusion mapping (dual sequence, fully quantitative) was implemented using Gadgetron^18^.

### Exercise echocardiography analysis

TTE post-processing analysis used EchoPac version 204 (GE Medical Systems). LA area was measured in the apical 4-chamber view in end-systole. Simpsons biplane ejection fraction was derived from the 4-chamber and 2-chamber views, by contouring the endocardial LV borders in end-diastole and end-systole. LV endocardial borders were manually traced at end-systole for strain analysis. LV rotation and rotation rates at the basal or apical short-axis planes were determined as average angular displacement of six myocardial segments. Curves of the averaged basal and apical rotation/ twisting rate in six segments and LV twist/twisting rate were directly generated from EchoPac. The systolic duration was measured from the onset of the QRS to the aortic valve closure, and the diastolic duration was calculated from the RR interval and systolic duration. To account for variable heart rates between subjects or during exercise, the time values were normalised to the percentage of systolic/diastolic duration(%). Time to peak twist in systole, time to peak twist rate in systole and time to peak untwist in diastole (diastolic untwisting time) were calculated using this method as a percentage of total systole/diastole duration.

Global longitudinal strain (GLS) was calculated by defining the endo and epicardial borders in the apical 4-chamber, 2-chamber and 3-chamber views using feature tracking and defining aortic valve closure (AVC) on the 3-chamber view.

### CMR Analysis

CMRs analysis used CVI42 (Circle Cardiovascular Imaging, Calgary, Canada). For myocardial blood flow (MBF) map analysis, LV endo- and epicardial contours were applied using machine learning with human oversight using a 16-segment model (32 for subendocardial and epicardial flow), with human oversight as needed (e.g. when there was no systolic blood pool cavity)(19). LV volume analyses and LV maximum wall thickness (MWT) were performed using a validated machine learning algorithm^20, 21^. LGE was quantified using the full-width half-maximum (FWHM) technique with LGE expressed in grams and as a % of total myocardium. An apical aneurysm (>5mm) or micro-aneurysm (<5mm) was defined by the presence of an akinetic/dyskinetic motion, scarring and a non-obliterating apical cavity typically distal to an area of obliteration.

### Statistical Analysis

Statistical analysis was performed in SPSS (IBM SPSS statistic, Version 26.0). Normality of data was assessed on histograms and using Shaprio-Wilk test. Continuous data was presented as mean+-standard deviation if normally distributed, or median (interquartile ranges) otherwise, and compared across participant groups using independent Student *t*-test or Mann-Whitney-U test. Within group comparisons for normally distributed continuous data were performed using the paired *t*-test. Categorical data were presented as counts and percentage and compared using Chi-square test. Correlation was assessed with Pearson’s coefficient. A linear regression model was used to determine which factors were associated with percentage predicted peak VO_2_ (PP peak VO_2_) in patients with overt and relative ApHCM. Interaction analysis was performed to look for any interaction between apical subendocardial MBF with either age or sex in the association with PP peak VO_2._ Exposures consisting of rest and exercise TTE, CMR and CPEX parameters were separately tested for their association with the outcomes using univariate linear regression. Unique, clinically relevant covariates with a *P* value <0.10 on univariate analysis were then entered into final multivariable regression models that adjusted for age and sex in PP peak VO2 and for MWT in subendocardial apical MBF models, using a forward stepwise procedure. The incremental value between steps was measured by the χ2 method. Multicollinearity of final model variables was excluded by confirming a variance inflation factor <3. A 2-sided *P*-value ≤0.05 was considered significant.

## RESULTS

A total of 46 subjects across the phenotypic spectrum of ApHCM were recruited for exercise transthoracic echocardiography with CPEX, and CMR. Four subjects were excluded (one each for: severe hypertension; knee pain precluding exercise; inadequate image quality and HCM misclassification). Of the remaining 42 subjects (age 54.1±12 years, 81% male, BSA 2.01±0.2m^2^), 27 met criteria (MWT ≥15mm) for overt ApHCM (age 56.7±11 years, 74% male, BSA 2.03±0.2m2) and 16 met criteria (MWT <15mm) for relative ApHCM (age 50.5±14 years, 94% male, BSA 1.94±0.2m^2^), **Figure 1**. They were compared with 36 HV subjects (age 50.0±8 years, 61% male, BSA 1.91±0.2m^2^), **Table 1**.

**Figure 1.**
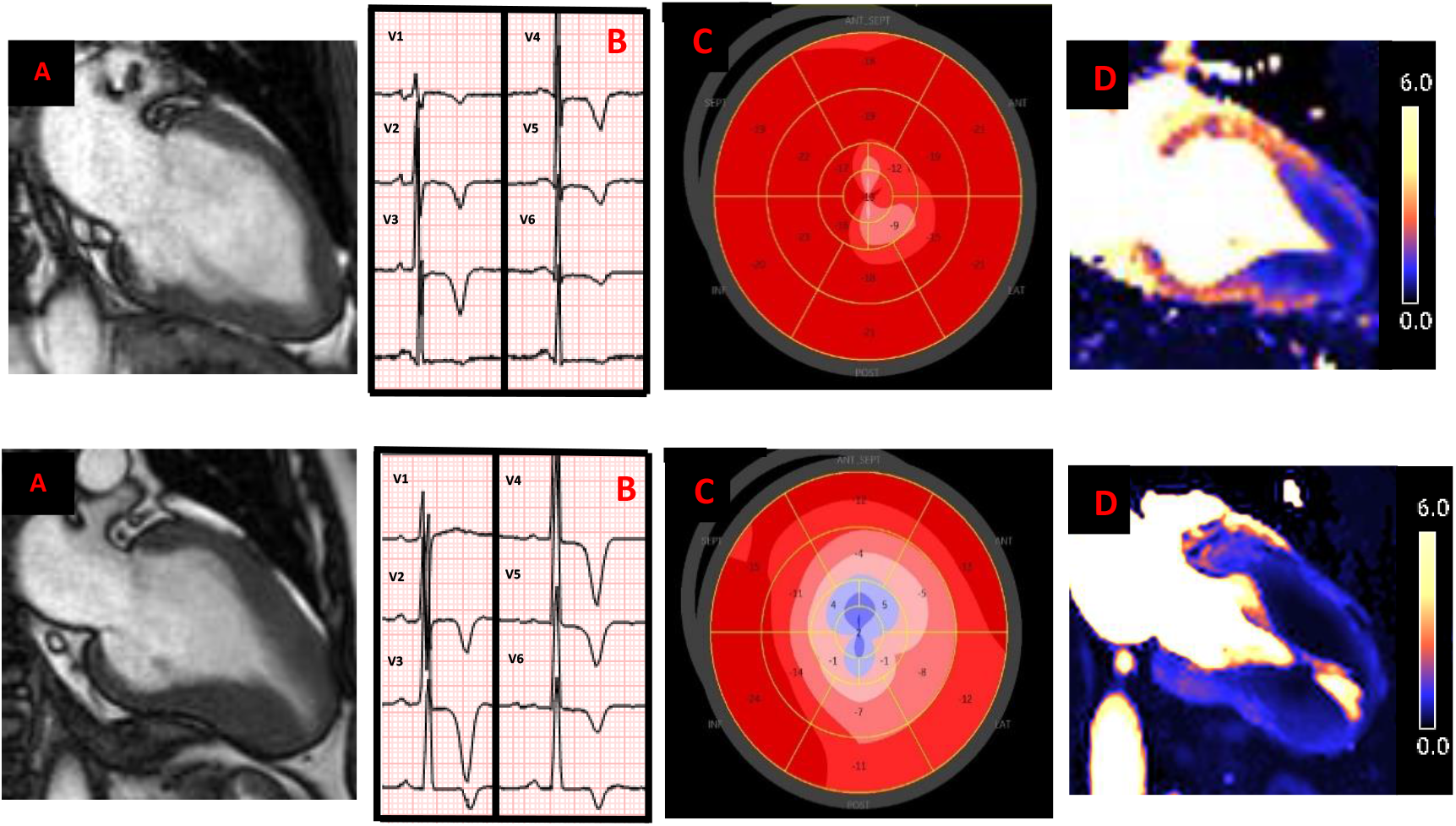
Comparison of relative and overt apical hypertrophic cardiomyopathy. Relative apical hypertrophic cardiomyopathy (ApHCM; top line) showing 2-chamber CMR cine in end-diastole demonstrating loss of apical tapering and mild hypertrophy (A). Electrocardiogram (ECG; B) showing precordial tall R-waves and deep T-wave inversion, global longitudinal strain (GLS) bullseye plot (C) demonstrating focal but mild apical impaired strain and profound apical perfusion defect on CMR stress perfusion mapping (D). Overt ApHCM (bottom line) showing unequivocal apical hypertrophy and characteristic ‘ace of spades’ appearance of left ventricular cavity (A). Similar ECG appearance to relative ApHCM but this time with giant negative T-waves in the precordial leads (B). GLS bullseye (C) demonstrating paradoxical apical strain and overall greater impairment in GLS and large perfusion defect, worse in the apex but with basal extension (D).

**Table 1.** Baseline characteristics of apical hypertrophic cardiomyopathy vs healthy controls.

|  | **Healthy**<br>**volunteers** | **Apical HCM** | **P value** |
| --- | --- | --- | --- |
| **N** | 36 | 42 |  |
| **Age** | 50.0±8 | 54.1±12 | 0.093 |
| **N (%) male** | 22 (61) | 34 (81) | 0.052 |
| **BSA (m<sup>2</sup>)** | 1.91±0.2 | 2.01±0.2 | 0.086 |
| **Diabetes, n (%)** | 1 (3) | 5 (12) | 0.132 |
| **Hypertension, n (%)** | 0 (0) | 13 (31) | **<0.001** |
| **Hypercholesterolaemia, n (%)** | 0 (0) | 8 (19) | **0.006** |
| **Indexed LA area (cm/m<sup>2</sup>)** | 9.1 (7-10) | 11.6 (10-14) | **<0.001** |
| **LV EDVi (ml/m<sup>2</sup>)** | 61.5±13 | 49.0±15 | **<0.005** |
| **LV ESVi (ml/m<sup>2</sup>)** | 20.9±7 | 14.0±6 | **<0.001** |
| **LVEF(%)** | 66.4±6 | 71.3±7 | **<0.005** |
| **MWT (mm)** | 9.8±2 | 17.5±5 | **<0.001** |
| **Apical aneurysm/**<br>**microaneurysm, n (%)** | 0 (0) | 8 (19) | **<0.001** |
| **Presence of apical LGE, n (%)** | - | 26 (66.7) | - |
| **LGE (% of total myocardium)** | - | 0.0 (8-18) | - |
| **Septal S'** | 7 (6,8) | 5 (4,6) | **<0.001** |
| **Septal E'** | 8 (7, 10) | 5 (3, 6) | **<0.001** |
| **Lateral S'** | 8 (6,9) | 5 (4,7) | **<0.001** |
| **Lateral E'** | 11±3 | 6.5±3 | **<0.001** |
| **LVOT gradient (mmHg)** | 6 (4, 7) | 6 (5, 9) | 0.130 |
| **Mid LV cavity gradient (mmHg)** | 0 (0, 0) | 0 (0, 20) | **<0.001** |
| **TR gradient (mmHg)** | 0 (0, 16) | 0 (0, 0) | **<0.005** |
| **Exercise septal S'** | 9 (8, 11) | 6 (5, 8) | **<0.001** |
| **Exercise septal E'** | 11.2±2 | 6.3±2 | **<0.001** |
| **Exercise lateral S'** | 9 (7, 10) | 6 (5, 8) | **<0.001** |
| **Exercise lateral E'** | 14 (11, 15) | 9 (6, 12) | **<0.001** |

Data was complete apart from no GLS (n=3, due to AF); no exercise data (n=1, exercise terminated early at patient request), no CMR data (n=1, claustrophobic). 3/42 ApHCM subjects had calculable longitudinal but not circumferential strain measures. In 14/42 subjects, resting strain parameters were calculable, but circumferential strain on exercise was not.

Comorbidities (diabetes, hypertension and hypercholesterolaemia) were present in the minority of ApHCM patients but, by definition, were absent in healthy volunteers (Table 1). Hypertension was more prevalent in overt than relative ApHCM (11/26(42%) vs 2/16(13%), p=0.042), but prevalence of diabetes and hypercholesterolaemia were equivalent (4/26(15%) vs 1/16(6%), p=0.375 and 4/26(15%) vs 4/16(25%), p=0.441 respectively).

### Baseline characteristics

Baseline characteristics are detailed in **Table 1**. ApHCM patients had smaller indexed LV end-diastolic and end-systolic volumes, higher ejection fractions and indexed left atrial areas. 26/41(63.4%) had apical scar. Apical aneurysms/microaneurysms were present in 8/42(19%). As expected, MWT was higher in the disease population (17.0±0.5mm vs 9.8±2mm, P<0.001). Septal and lateral S’ and E’ measures were lower both at rest, and during exercise in subjects with ApHCM (P<0.001 for all), and inversely correlated with indexed LA area. LVEF was equivalent between overt and relative ApHCM, and as expected, MWT was lower in the relative subgroup (12.8±1mm vs 19.6±5mm, P<0.001). Aside from septal S’ at rest, septal and lateral S’ and E’ measures were higher in the relative subgroup at rest and exercise.

### Apical hypertrophic cardiomyopathy strain

#### Rest strain

**Table 2** demonstrates rest and exercise strain. Both global and regional (apical) longitudinal strain were markedly reduced in ApHCM compared with controls (−11.0%(−15,−7) vs −18.3%(−20,−17) and −8.6±7% vs −20.4±4% respectively, P<0.001 for both). The degree of overall LV rotation in ApHCM was greater (22.6±9⸰ vs 16.6±4⸰, P<0.005), with LV twist and untwist rates equivalent, but time to peak LV twist occurred earlier in systole (41.7±10% vs 49.8±15%, P=0.011) and LV untwist occurred later in diastole (17.9% (12,26) vs 9.2% (5,17), P<0.005 (**Figure 2**).

**Figure 2.**
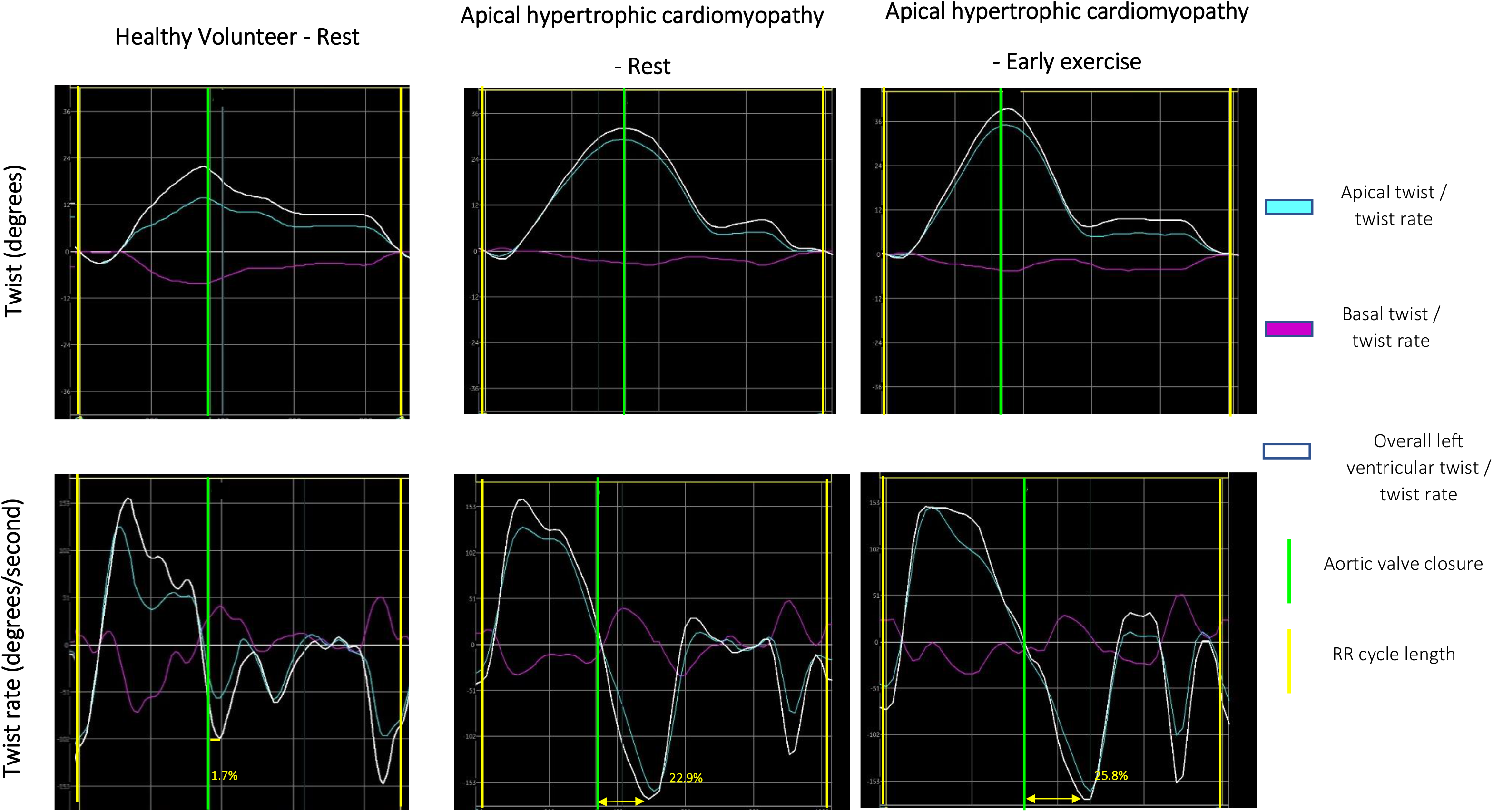
Twist and twist rate comparison of healthy volunteer (HV) and subject with apical hypertrophic cardiomyopathy (ApHCM) at rest and on exercise. Visually, it is easy to appreciate the increased apical and left ventricular (LV) twist in ApHCM compared with HV, which augments further on exercise. The peak apical and LV twist in HV is 4.8⸰ and 9.4⸰ respectively. In ApHCM, at rest it is 25.4⸰ and 30.1⸰ which increases to 35.4⸰ and 38.5⸰ on exercise. The time to peak LV untwist is much shorter in the HV (1.7%) compared with ApHCM at rest (22.9%), increasing to 25.8% on exercise.

**Table 2.** Comparison of longitudinal strain and twist parameters performed at rest between subjects with apical hypertrophic cardiomyopathy (ApHCM) and healthy volunteer (HV) controls and comparing rest and exercise in ApHCM.

|  | **HV**<br>**Rest** | **ApHCM rest** | **ApHCM early**<br>**exercise** | **P value**<br>**(HV vs**<br>**ApHCM rest)** | **P value (rest**<br>**vs exercise**<br>**ApHCM)** |
| --- | --- | --- | --- | --- | --- |
| **GLS (%)** | -18.3 (-20, -17) | -11.0 (-15, -7) | -12.5 (-16, -9) | **<0.001** | 0.201 |
| **Apical longitudinal strain (%)** | -20.4±4 | -8.6±7 | -10.9±8 | **<0.001** | **0.011** |
| **Peak apical rotation (°)** | 10.9±4 | 15.7±9 | 23.0±7 | **0.013** | **0.007** |
| **Peak basal rotation (°)** | -6.7±3 | -5.9 (-9, -3) | -6.5 (-11, -5) | 0.924 | 0.308 |
| **Peak LV rotation (°)** | 16.6±4 | 22.6±9 | 30.6±7 | **<0.005** | **0.006** |
| **Time to peak LV rotation (%)** | 90.2±10 | 91.4±9 | 89.2±13 | 0.622 | 0.248 |
| **Peak apical twisting rate (°/s)** | 72.1 (53, 86) | 94.1 (67, 128) | 164.9 (127, 193) | **0.017** | **<0.001** |
| **Peak basal twisting rate (°/s)** | -72.2 (-86, -55) | -52.3 (-92, -37) | -91.5 (-137, -59) | 0.081 | **<0.005** |
| **Peak LV twisting rate (°/s)** | 106.3 (90, 121) | 126.3 (81, 160) | 213.9 (166, 236) | 0.143 | **<0.001** |
| **Time to peak LV twisting rate (%)** | 49.8±15 | 41.7±10 | 40.0±13 | **0.011** | 0.657 |
| **Peak apical untwisting rate (°/s)** | -60.2 (-94, -50) | -64.5 (-87, -48) | -140.6 (-167, -101) | 0.901 | **<0.001** |
| **Peak basal untwisting rate (°/s)** | 50.3 (36, 68) | 46.1 (37, 61) | 57.7 (37, 120) | 0.858 | 0.090 |
| **Peak LV untwisting rate (°/s)** | -99.6 (-117, -75) | -98.4 (-144.6, -73.9) | -177.4 (-249, -129) | 0.408 | **<0.001** |
| **Time to peak LV untwisting rate (%)** | 9.2 (5, 17) | 17.9 (12, 26) | 25.3 (15-34) | **<0.005** | 0.070 |

Regional assessment of apical twist showed that the magnitude of apical rotation was greater (15.7±9⸰ vs 10.9±4⸰, P=0.013) and faster (apical twisting rate 94.1⸰/s (67,128) vs 72.1⸰/s (53,86), P=0.017) in ApHCM, but the rate of untwist similar to controls. Basal rotation, twist rate and untwist rate were equivalent. Regional results suggest that overall LV twist and untwist is largely driven by apical, rather than basal changes.

Comparison of ApHCM subtypes showed graded severity of longitudinal strain, with lesser impairment in relative than overt ApHCM (global and apical), but worse than controls, **figure 1** (an example) and **supplementary table 1**. Twist was equivalent, but diastolic untwist time was less delayed in relative ApHCM.

#### Exercise strain

In ApHCM, global and apical longitudinal strain increased during exercise. Measures of twist and untwist were augmented globally and apically for rotation (P=0.005; P=0.007), as were twist rate (both P<0.001) and untwist rate (both P<0.001). However, diastolic untwisting time, which was delayed at rest (18% into diastole compared to 9% for controls), became numerically more delayed (25%) during exercise, although this failed to reach significance (P=0.070) (**Table 2**, **Figure 2**).

#### Functional capacity

Peak VO_2_ was variable and lower in ApHCM than controls (23.4±8ml/kg/min vs 30.7±7ml/kg/min, P<0.001) with 35% overall having reduced (<80% predicted) peak VO_2_ (5/16 (31%) relative ApHCM and 9/26 (35% overt ApHCM) vs 6% of controls (P<0.005). There was no difference in prevalence of comorbidities with or without functional limitation (diabetes: 3/14(21%) with PP peak VO_2_ <80% vs 2/28(7%), p=0.210 without; hypertension 5/14(36%) vs 8/28(29%), p=0.750, hypercholesterolaemia (4/14(29%) vs 4/28(14%), p=0.320). Other CPEX markers were equivalent to controls (VE/VCO2 slope, P=0.051; OUES, P=0.236, O_2_ pulse, P=0.566).

### Myocardial mechanics, blood flow and functional capacity

Multiple parameters of myocardial function appeared independent: LVEF had no association with global (or apical) longitudinal strain, which itself had no association with any twist/untwist parameters at rest or during exercise. Yet there were structural and mechanical associations: GLS worsened with increased apical hypertrophy (r= 0.603, P<0.001) and with markers of impaired LV filling; septal E’ (r= −0.421, P=0.015), and lateral E’ (r= −0.573, P<0.001). There were also structural and myocardial blood flow associations; lower apical subendocardial MBF was associated with (i) more impaired long axis function in systole and diastole (lower septal E’ (r= −0.347, P=0.041), lateral S’ (r=0.465, P=0.005) and lateral E’ (r=0.517, P<0.001)), (ii) worsening (higher) GLS (r= −0.621, P<0.001), (iii) apical longitudinal strain (r= −0.567, P<0.001), and (iv) a longer diastolic untwisting time (r=-0.350, P=0.039). All associations were present at rest but were more closely associated on exercise.

The PP peak VO_2_ considers age, sex and body size, and its associations were explored using a univariate analysis (**Supplementary Table 2)**. PP peak VO_2_ was worse with lower lateral S’ at rest (P=0.046), and more impaired GLS both at rest (p=0.017) and exercise (P=0.048). It was also lower with longer diastolic untwisting time on exercise (P=0.011) and apical subendocardial MBF (P<0.005). On multivariate analysis, the only independent association with PP peak VO_2_ was diastolic untwisting time, when controlled for age and sex (**Supplementary Table 3,** R^2^ for model 0.469, P=0.030). There were no interactions between apical subendocardial MBF and age or sex in the association with PP peak VO_2_.

ApHCM subjects with apical aneurysms (5/8 microaneurysms, 3/8 aneurysms; all overt ApHCM) had a greater amount of LGE and more impaired longitudinal strain than subjects without. Diastolic untwisting time was numerically (but not statistically) longer, but other baseline CMR characteristics and twist parameters were equivalent (**Tables 3a and 3b**). 2/8 aneurysm subjects had PP peak VO_2_ <80%.

**Table 3a.** Baseline imaging comparison between apical hypertrophic subjects with and without apical aneurysms.

|  | Aneurysm | No aneurysm | P value |
| --- | --- | --- | --- |
| **N** | 8 | 34 |  |
| **Age (years)** | 55.8±10.4 | 53.7±12.7 | 0.630 |
| **BSA (m<sup>2</sup>)** | 2.05±0.1 | 2.0±0.2 | 0.370 |
| **LA area indexed (cm/ m<sup>2</sup>)** | 11.1 (9.8, 12.2) | 11.6 (9.5, 13.9) | 0.550 |
| **LVEDVi (ml/ m<sup>2</sup>)** | 45.2±14.1 | 51.5±13 | 0.278 |
| **LVESVi (ml/ m<sup>2</sup>)** | 13.8±4.7 | 14.5±5.8 | 0.735 |
| **LVEF (%)** | 69.9±4.2 | 71.6±7.2 | 0.383 |
| **Global stress MBF (ml/g/min)** | 1.65±0.5 | 1.86±0.5 | 0.259 |
| **Apical stress MBF (ml/g/min)** | 1.16 (0.9, 1.28) | 1.46 (1.14, 1.93) | 0.097 |
| **LGE (%)** | 22.6 (17.6, 30.6) | 5.9 (0.0, 17.0) | **<0.005** |

**Table 3b.**
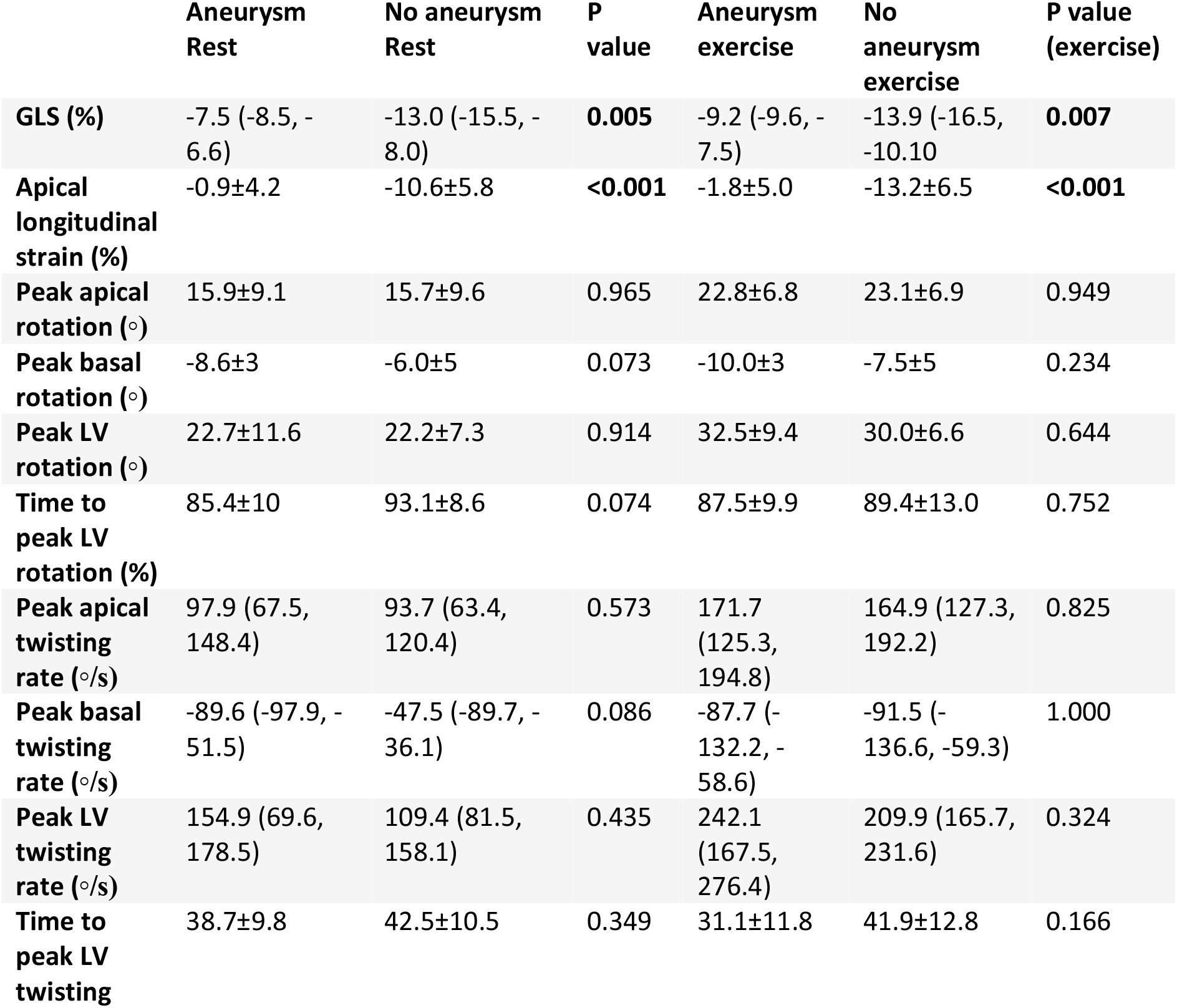
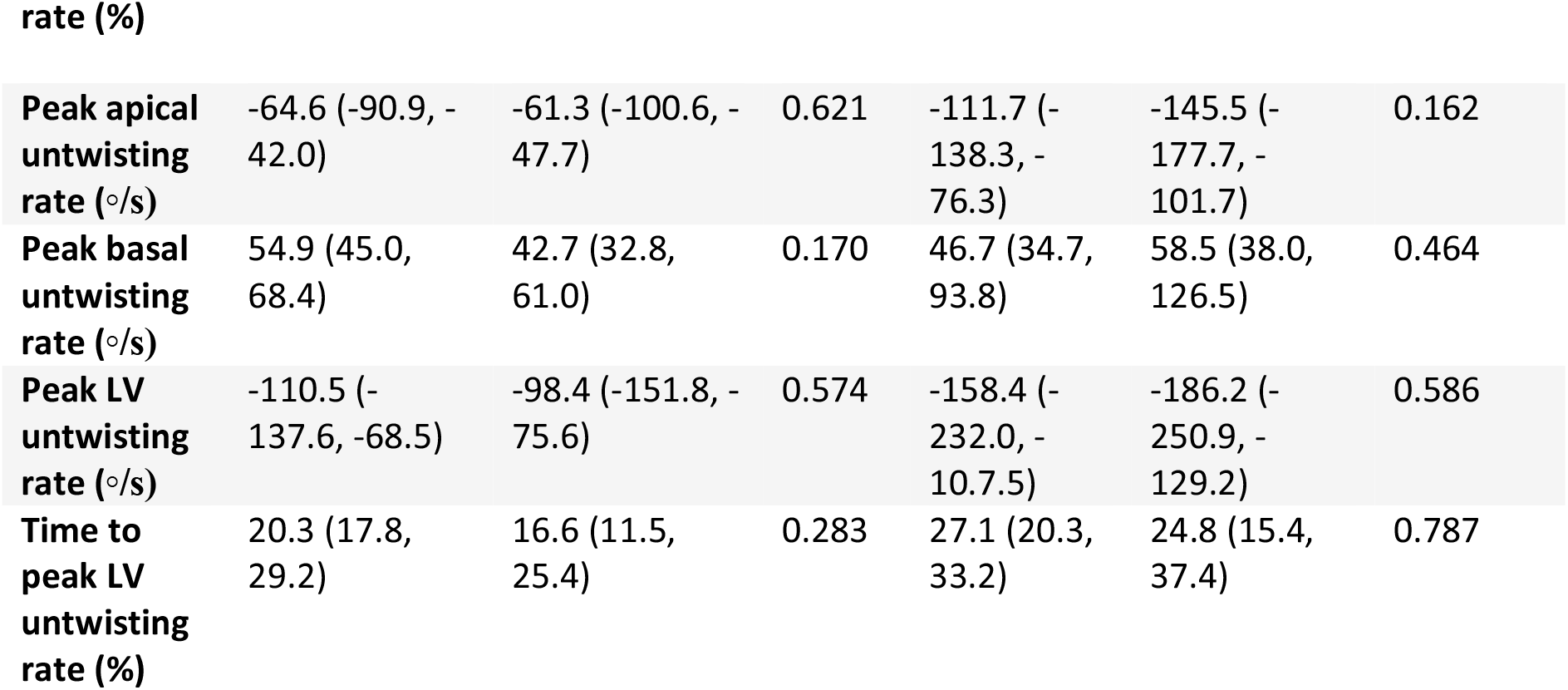
Comparison of strain parameters at rest and exercise in ApHCM subjects with and without apical aneurysms.

|  | Aneurysm<br>Rest | No aneurysm<br>Rest | P<br>value | Aneurysm<br>exercise | No<br>aneurysm<br>exercise | P value<br>(exercise) |
| --- | --- | --- | --- | --- | --- | --- |
| **GLS (%)** | -7.5 (-8.5, -6.6) | -13.0 (-15.5, -8.0) | **0.005** | -9.2 (-9.6, -7.5) | -13.9 (-16.5, -10.10) | **0.007** |
| **Apical longitudinal strain (%)** | -0.9±4.2 | -10.6±5.8 | **<0.001** | -1.8±5.0 | -13.2±6.5 | **<0.001** |
| **Peak apical rotation (°)** | 15.9±9.1 | 15.7±9.6 | 0.965 | 22.8±6.8 | 23.1±6.9 | 0.949 |
| **Peak basal rotation (°)** | -8.6±3 | -6.0±5 | 0.073 | -10.0±3 | -7.5±5 | 0.234 |
| **Peak LV rotation (°)** | 22.7±11.6 | 22.2±7.3 | 0.914 | 32.5±9.4 | 30.0±6.6 | 0.644 |
| **Time to peak LV rotation (%)** | 85.4±10 | 93.1±8.6 | 0.074 | 87.5±9.9 | 89.4±13.0 | 0.752 |
| **Peak apical twisting rate (°/s)** | 97.9 (67.5, 148.4) | 93.7 (63.4, 120.4) | 0.573 | 171.7 (125.3, 194.8) | 164.9 (127.3, 192.2) | 0.825 |
| **Peak basal twisting rate (°/s)** | -89.6 (-97.9, -51.5) | -47.5 (-89.7, -36.1) | 0.086 | -87.7 (-132.2, -58.6) | -91.5 (-136.6, -59.3) | 1.000 |
| **Peak LV twisting rate (°/s)** | 154.9 (69.6, 178.5) | 109.4 (81.5, 158.1) | 0.435 | 242.1 (167.5, 276.4) | 209.9 (165.7, 231.6) | 0.324 |
| **Time to peak LV twisting** | 38.7±9.8 | 42.5±10.5 | 0.349 | 31.1±11.8 | 41.9±12.8 | 0.166 |
| **Peak apical untwisting rate (°/s)** | -64.6 (-90.9, -42.0) | -61.3 (-100.6, -47.7) | 0.621 | -111.7 (-138.3, -76.3) | -145.5 (-177.7, -101.7) | 0.162 |
| **Peak basal untwisting rate (°/s)** | 54.9 (45.0, 68.4) | 42.7 (32.8, 61.0) | 0.170 | 46.7 (34.7, 93.8) | 58.5 (38.0, 126.5) | 0.464 |
| **Peak LV untwisting rate (°/s)** | -110.5 (-137.6, -68.5) | -98.4 (-151.8, -75.6) | 0.574 | -158.4 (-232.0, -107.5) | -186.2 (-250.9, -129.2) | 0.586 |
| **Time to peak LV untwisting rate (%)** | 20.3 (17.8, 29.2) | 16.6 (11.5, 25.4) | 0.283 | 27.1 (20.3, 33.2) | 24.8 (15.4, 37.4) | 0.787 |

## DISCUSSION

Contrary to previous published results, our ApHCM cohort demonstrated a ‘gain in function’ of twist, with increased apical and LV rotation and increased rate of apical and LV twist. However, untwist the LV diastolic untwisting time was slowed, occurring later in diastole, and was exacerbated by exercise. We also demonstrated markedly impaired global and apical longitudinal strain, which improved with exercise. However, one of the most compelling findings was the demonstration that one third of ApHCM patients have functional limitation, which associated with mechanical and microvascular abnormalities (impaired GLS and apical MBF). Yet, functional limitation was best predicted by diastolic untwisting time. These results begin to provide a narrative as to the complex interlinked structural and functional mechanisms of this disease (**central illustration)**. We hypothesize that abnormalities of myocardial contraction/relaxation and MBF lead to diastolic dysfunction with a “positive feedback loop” between prolonged systole/shortened diastole reducing myocardial blood flow, which itself reduces the oxygen supply needed for active myocardial relaxation, with a net clinical effect of functional limitation.

LV untwist plays a pivotal role in cardiac contractility as it helps generate suction power in early diastole^22, 23^. Delayed untwisting despite normal or increased LV twist has been observed in states associated with impaired relaxation, such as pressure-loading^24^ and HCM^11, 14, 25^, whereas impaired twist has mainly been shown in cardiomyopathies with impaired pump function^10, 26^. Mitral annular velocity (E’; an index of rapid ventricular filling measurements) is an excellent marker of LV relaxation^27^ and is impaired in ApHCM both at rest and on exercise. Our findings support that despite a gain in circumferential twist function, longitudinal function and diastolic relaxation are impaired.

The subendocardium is the region most vulnerable to ischemia and increased wall stress^28^,and predominantly affects longitudinal shortening function^10^. Here we demonstrate a strong association between impaired apical MBF (most impaired subendocardially) and GLS, providing evidence of a potential link between microvascular ischemia and contractile dysfunction. Lower apical MBF was also associated with a longer diastolic untwisting time. Stephenson et al. previously associated ‘contractile persistence’ with a greater impairment in myocardial perfusion reserve in ApHCM patients with chest pain. It is difficult to differentiate prolonged contraction from delayed relaxation, although arguably they have the same net effect. In the hypertrophied apex, the reduction in diastolic coronary blood flow can be explained by delayed and impaired relaxation and increased filling pressures causing extravascular compression. Plausibly, the ischemia itself may also accentuate delayed relaxation by causing myocyte damage, creating a feedback loop of altered myocardial mechanics and ischemia^29^. The clinical effect is of limitation, supporting a previous finding in HCM that peak VO_2_ is associated with coronary flow reserve^29^. On a cellular level, recurrent ischemia is likely to result in myocyte death and replacement fibrosis, further attenuating GLS (and eventually overall systolic function, which is seen in the ‘burn-out’ phase), which relates to fibrosis in HCM^29^. Unlike non-apical HCM, whereby GLS improves but twist does not^25^, we’ve shown overall improved contractile response to exercise, but at the expense of diastolic relaxation, which is likely driven by ischemia.

We postulate that apical rotation is greater and faster than controls because, as is known in HCM, there is an energetic ‘gain in function’ within the sarcomere resulting in hypercontractility due to excessive actin-myosin cross-bridges. Circumferential ‘twist’ is mostly controlled by subepicardial fibres, which are often the last to be affected by myocardial diseases. Usually, by the time the mid and subepicardium are affected by disease, as described by Modesto et al, a reduction in twist is also seen. Therefore, reduced longitudinal strain despite normal or supranormal ejection fraction reflects subendocardial dysfunction, but without affecting the subepicardial layers, resulting in increased twist and twist-rate (**Figure 3**).

**Figure 3.**
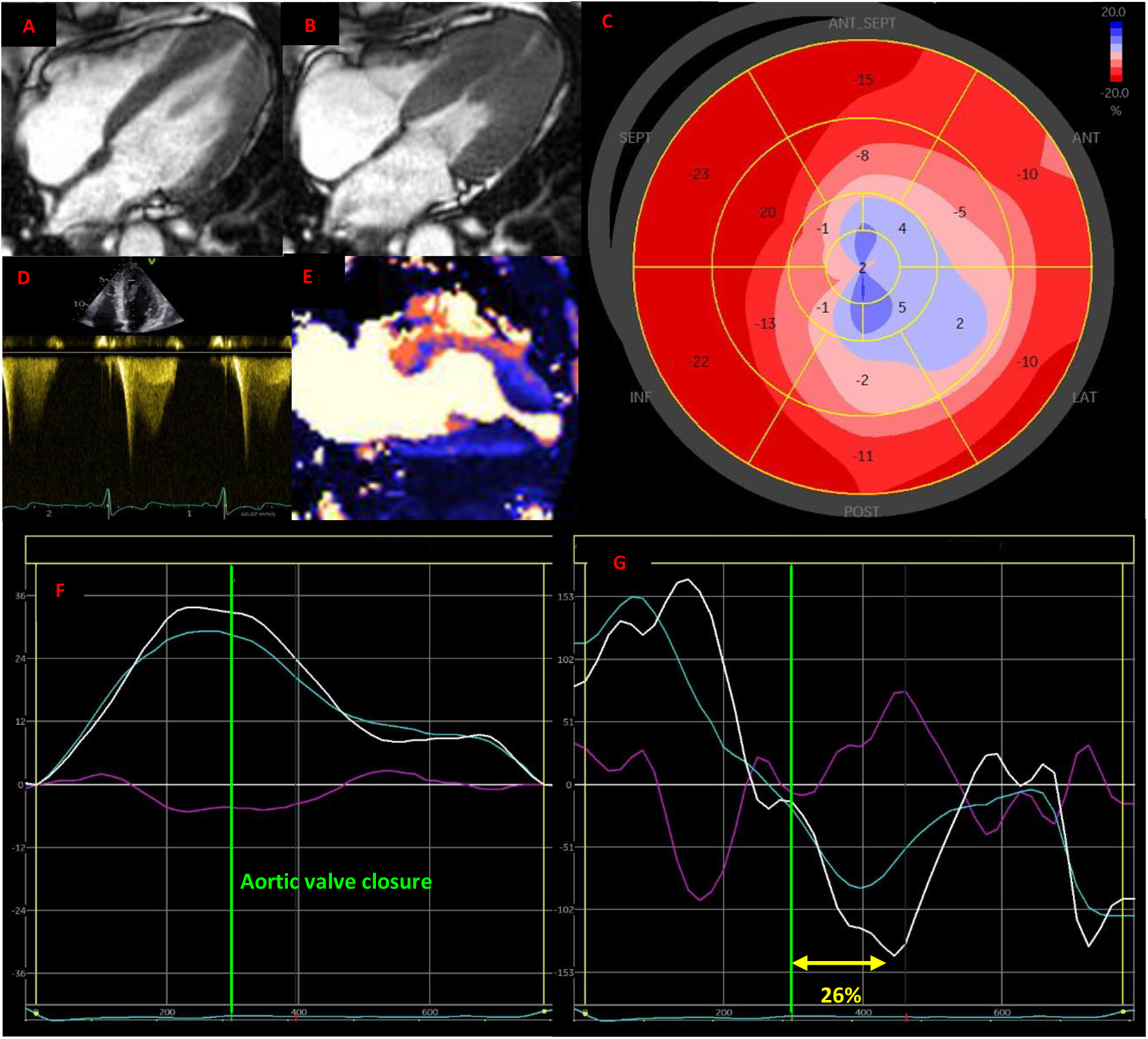
Multiparametric transthoracic echocardiographic (TTE) and cardiac magnetic resonance (CMR) imaging findings in a patient with overt apical hypertrophic cardiomyopathy and an apical aneurysm. **A.** CMR 4-chamber image in end-diastole demonstrating apical hypertrophy. **B.** CMR 4-chamber image in end-systole showing presence of apical aneurysm with cavity obliteration proximal to aneurysm. **C.** Global longitudinal strain bullseye plot showing reduced and paradoxical apical strain typical of an apical aneurysm. **D.** Continuous wave doppler trace through mid-left ventricle (LV) on TTE showing classic doppler appearance of an apical aneurysm with early systolic peak and subsequent signal drop-out and a distinct paradoxical diastolic flow jet. **E.** 2-chamber stress perfusion CMR map demonstrating impaired apical perfusion, most notable in the subendocardium. **F.** TTE twist graph showing increased apical and LV twist. **G.** TTE twist rate graph demonstrating increased rate of apical and LV twist, but most notably, delayed rate of untwist in early diastole at 26%.

Lastly, we have demonstrated that exercise capacity is not associated with MWT, highlighting that although the diagnosis of ApHCM is principally based on identifying apical hypertrophy, mechanistically, there are other pathological processes that cause morphological, mechanical, electrocardiographic, and functional impairment. Combined with the observed reduction in GLS, large gain in twist function and delayed untwist adds credence to the assertion that relative ApHCM is indeed a mild or ‘early’ disease phenotype, and that it should be treated as such, highlighting the need to revisit the diagnostic criteria for ApHCM.

We propose that our results differ from previous ApHCM publications as our patient cohort is considerably larger and more diverse, including a high proportion with relative ApHCM, who we have demonstrated have the largest ‘gain in function’ of twist parameters.

## LIMITATIONS

Exercise twist parameters were not obtained in our control group. Other measures of diastolic function (peak early (E) and late (A) diastolic velocities of mitral inflow) were not acquired. Few patients had apical aneurysms, preventing subgroup analyses. The protocol was optimised for ventricular, not atrial assessment, therefore LA volumes could not be derived, only LA area. Echocardiographic strain was performed over CMR strain as current CMR analysis techniques were found unreliable blood pool was absent (here with apical cavity systolic obliteration). ApHCM subjects were recruited from tertiary cardiomyopathy clinics therefore potential exists for selection bias. Of those initially approached for recruitment, some subjects undergoing CMR did not undergo CPEX exercise echocardiogram. This was due to scheduling difficulties precluding a return for CPEX (n=3), unsuitable for exercise due to mobility (n=6) or declined CPEX (n=3).

## CONCLUSION

One third of ApHCM patients have functional limitation, independent of the degree of apical hypertrophy. All ApHCM patients demonstrate LV mechanical dysfunction (increased twist, delayed diastolic untwist, impaired longitudinal strain), as well as apical perfusion defects suggestive of microvascular ischemia. The degree of impairment in apical MBF is associated with delayed diastolic untwist, which is an independent predictor of exercise capacity. We propose that the delay in diastolic relaxation is the unifying feature linking mechanical, functional and physiological impairment in ApHCM by interfering with normal MBF in diastole, contributing to apical microvascular ischemia and reduced exercise capacity.

## SUPPLEMENTARY MATERIAL

Supplementary Tables 1-3

## Data Availability

Data can be made available upon reasonable request

## ABBREVIATIONS

ApHCM: apical hypertrophic cardiomyopathy
AVC: aortic valve closure
CMR: cardiac magnetic resonance
CPEX: cardiopulmonary exercise testing
ECG: electrocardiogram
GLS: global longitudinal strain
HV: healthy volunteer
LGE: late gadolinium enhancement
LV: left ventricle/ventricular
MWT: maximum wall thickness
PP peak VO_2_: percentage predicted peak oxygen uptake
TTE: transthoracic echocardiography

